# Evaluation of a One Health public health program based on minimum inputs to control *Taenia solium* in Madagascar

**DOI:** 10.1101/2024.08.30.24312828

**Authors:** Diana Edithe Andria-Mananjara, Modestine Raliniaina, Mihaja Rakotoarinoro, José A Nely, Nivohanitra Razafindraibe, Sylvia Noromanana Ramiandrasoa, Betthelhein Ramahefasoa, Valisoa Claude Rakotoarison, Paul R. Torgerson, Eric Cardinale, Harena Rasamoelina-Andriamanivo, Glenn Edosoa, Agnès Fleury, Kabemba E. Mwape, Bernadette Abela, Marshall W. Lightowlers, Meritxell Donadeu

## Abstract

Cysticercosis in humans caused by the parasite *Taenia solium* is one of the World Health Organization’s Neglected Tropical Diseases. The parasite is transmitted between the human host and pigs. Efforts to prevent the disease have relied mainly on treatment of people with anthelmintics. However to date there is no practical and effective control method that has been delivered as a public health program. Here we describe a large-scale, minimum inputs *T. solium* control program implemented as a public health program in Madagascar. Initially pigs were vaccinated for porcine cysticercosis and medicated with oxfendazole, after which only young piglets and pigs imported into the program area were targeted for interventions. After piglet interventions were in place and on-going, a single mass drug administration (MDA) was delivered to the human population with a taeniacide. The outcomes were assessed one year after the human treatment, by comparing pre-and post-intervention levels of human *T. solium* taeniasis and porcine cysticercosis caused by *T. solium*. Over a twenty-two-month period, 96,735 pig vaccinations were delivered and during the MDA, 117,216 people received taeniacide. Ninety percent of the pig population were receiving vaccination and medication at the end of the intervention period. Coverage of the eligible human population by the MDA was 62.5%. Human taeniasis was found to be 1.25% prior to the MDA and 0.6% one year after the MDA. Prior to the intervention 30.8% of slaughter-age pigs had viable *T. solium* infection whereas no viable infection was detected in any pig treated in the program. The program successfully demonstrated effective control of *T. solium* transmission using minimum inputs and delivered as a public health program. Sustained control and expansion of the program could potentially lead to the elimination of the disease being a public health problem in Madagascar.

**Author summary:** Cysticercosis caused by the cestode parasite *Taenia solium* is one of the World Health Organization’s neglected tropical diseases. We implemented One Health cysticercosis control as a public health program in a highly endemic region of Madagascar. The program was designed to require minimum inputs while achieving maximum impacts on control within a short period of time. Pigs were vaccinated with the TSOL18 vaccine and simultaneously treated with oxfendazole. Over a 2-year period, piglets 2-3 months of age were treated. After pig vaccination practices where established, the human population received a single treatment with a taeniacide. The program was evaluated by assessment of porcine cysticercosis and human taeniasis before and 12 months after the human mass drug administration. More than 95,000 pig vaccinations were delivered and 117,000 persons received taeniacide. The intervention entirely eliminated viable porcine cysticercosis in slaughter-age pigs that had received treatment as well as reducing human taeniasis. Continuation and expansion of the program would have the potential to eliminate *T. solium* from the program area, or even from the entire country.

## Introduction

Neurocysticercosis (NCC) is an infection of the central nervous system caused by the larval stage of a zoonotic parasite, *Taenia solium* [1]. It is prevalent in many parts of the world where risk factors for the parasite’s life cycle are present, such as poverty, lack of hygiene and sanitation, and the presence of free-roaming pigs [1]. As a result, many low and middle income countries, including Madagascar, are endemic to *T. solium* [2, 3], and NCC is a leading cause of epilepsy in the population [4].

NCC is considered a preventable and potentially eradicable neglected tropical disease [5]. So far, attempts to control transmission of *T. solium* have relied mainly on preventive chemotherapy in humans using either praziquantel or niclosamide [6–9]. Another tool available for the control of *T. solium* is the combined use of the TSOL18 (Cysvax®) porcine cysticercosis vaccine together with a single treatment with oxfendazole in pigs. The vaccine has proved to be safe and efficient, and the combination with oxfendazole has been found to eliminate parasite transmission by the treated pigs in field trials undertaken in Nepal, Cameroon and Uganda [10–12].

One Health research projects based on mass drug administration (MDA) and/or targeted chemotherapy in combination with other One Health measures (mainly pig vaccination and/or treatment) have achieved a reduction in *T. solium* transmission [13–17]. However, the approaches used have been neither practical, sustainable nor able to be implemented successfully as public health programs. There are currently no validated models for the control of the parasite.

Here we sought to evaluate the effectiveness of a *T. solium* control strategy requiring minimum inputs with the potential to lead to a substantial and sustainable reduction in parasite transmission, and delivered as a public health program by the Veterinary Services of the Ministry of Livestock and the Ministry of Public Health of Madagascar. Principal components of the program involved vaccination and treatment of young piglets, and a single round of MDA in the human population after pig vaccination had been well established. Outcomes were assessed by comparing pre and post intervention levels of human *T. solium* taeniasis and porcine cysticercosis.

## Methods

### Ethics statement

Intervention and assessment protocols were designed and implemented in compliance with ethical approvals for the study granted by the Madagascar Ministry for Public Health Ethics Committee for Biomedical Research No. 088-MSANP/SG/AMM/CERBM and by the Malagasy National Animal Ethics Committee, No. 001-21/CENA.

### Study area and timeline

The control program was implemented from August 2021 to August 2023 in a single contiguous area in central Madagascar comprising nine municipalities in two districts of Vakinankaratra region, Betafo and Mandoto (Fig 1). The area comprises approximately 420 villages (hamlets) spread over 84 fokontany (the lowest administrative unit), 205,197 inhabitants in 45,539 households [18] and an estimated 27,000 pigs. The most important economic activity among 93% of the population is mixed cropping with livestock farming [18]. Pig farming is one the major livestock activities of households. It is used as a means of livelihood and a source of savings for small scale farmers [19]. The program area was selected because the prevalence of human cysticercosis in the region was relatively high [20]. It is also one of the main suppliers of pigs for Antsirabe (the largest city in the region) and the capital of Madagascar, Antananarivo [21], and cases of porcine cysticercosis were seen frequently in Vakinankaratra compared to other regions of the central highlands [22].

**Fig 1.**
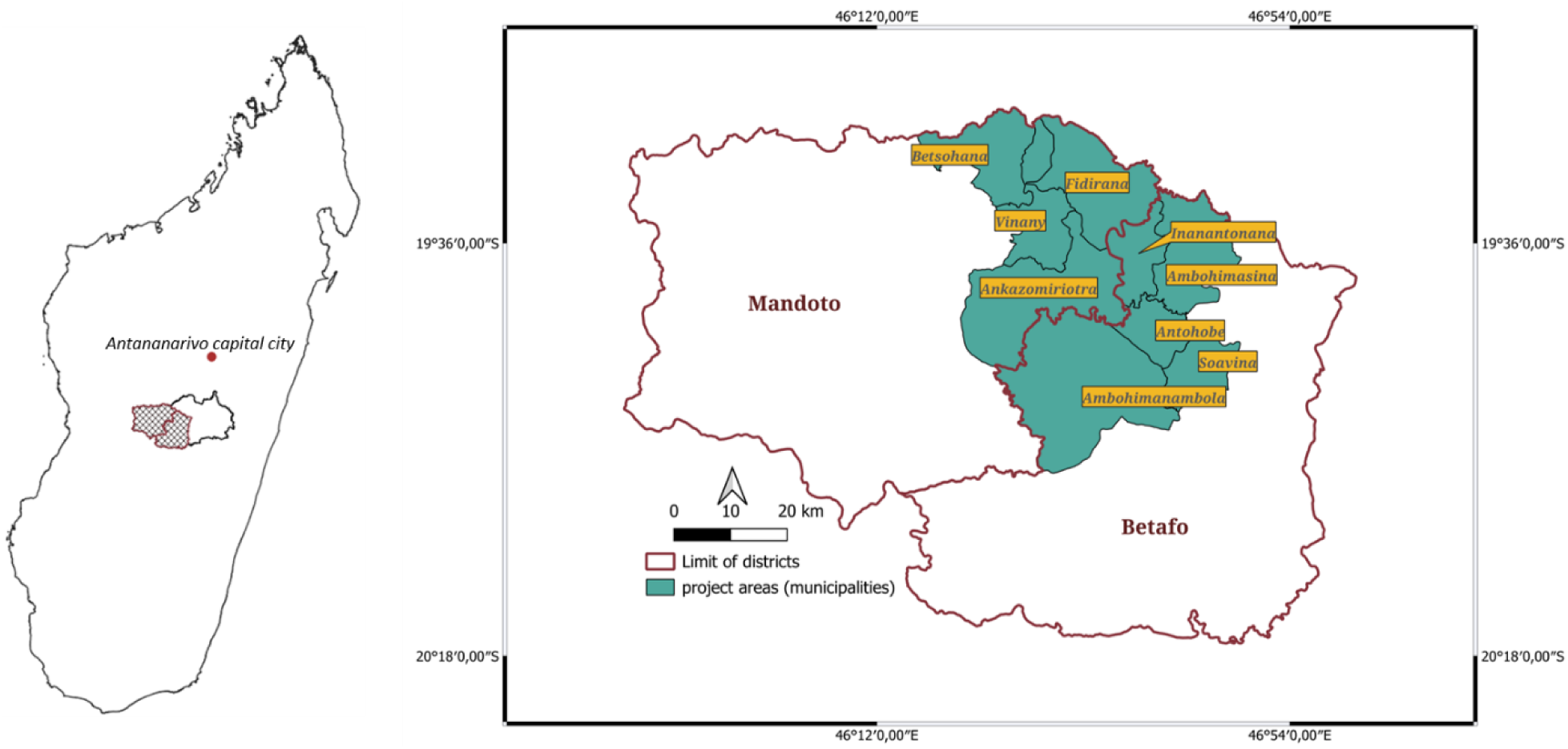
*Taenia solium* One Health control program implementation areas in Madagascar

### Program design and rationale

Interventions to interrupt *T. solium* transmission were undertaken in the two obligate hosts of the parasite, pigs and humans. Implementations were designed as One Health interdependent actions and included training of actors, advocacy and social mobilization to maximize the involvement of the relevant stakeholders.

The timings of the interventions were a key component of the program design. Because humans after receiving the MDA can get infected or re-infected with *T. solium* after ingesting contaminated pork (as there is no immunity to the tapeworm), the first step was to reduce the risk of infection in humans by reducing the prevalence of the disease in the pig population. Therefore, the first step was to vaccinate and treat the pigs to minimise the risk of infection or reinfection in humans. Only when that risk was minimised by having the pig population vaccinated and treated, then the MDA was to be conducted. However, because of the likely imperfect coverage of the MDA, and the potential risk of tapeworm eggs surviving in the environment, the vaccination of piglets and incoming pigs continued for a year after the MDA.

While the implementation was based on activities conducted as a public health program (therefore implemented by existing staff linked to the Ministry of Public Health or the Veterinary Services), the evaluation was conducted as a research project so as to obtain accurate measures of the impact of the intervention on *T. solium* transmission. The local coordination and supervision were provided by the National Center for Applied Research on Rural Development (FOFIFA).

### Pig intervention

Intervention activities in pigs were undertaken between October 2021 and August 2023. Initially a census of the pig population in the program areas was conducted to determine the number of pigs. Subsequently, pigs in the area were vaccinated with the TSOL18 vaccine (Cysvax® from Indian Immunologicals Limited) and, simultaneously, received an oral treatment with oxfendazole (Oxfenvet® from Indian Immunologicals Limited). Interventions in the pig population were undertaken by the staff of the Ministry of Livestock through the Veterinary Services Directorate, the sanitary veterinarians and their animal health agents (vaccinators).

At the start of the program, the intervention targeted all the pigs in the population. Inclusion criteria were animals ≥ 2 months of age, showing no obvious signs of sickness and not pregnant or lactating. The animals received two immunizations, approximately one month apart, with the Cysvax® vaccine and, at the time of the second immunization they also received treatment with oxfendazole at 30 mg/kg. Subsequently, throughout the program, newborn piglets (at ≥ 2 months of age) and any pigs imported into the program area received both Cysvax vaccination and oxfendazole treatment twice, at least 1 month apart (Fig 2). At the time of the first vaccination and treatment, the animals were tagged with a button tag in one ear. At the time of the second vaccination and treatment, the button tag was marked with indelible pen. Vaccinators visited the villages approximately on a monthly basis, and farmers having newborn piglets or imported animals were identified by the farmer themselves, the Chief of the fokontany or neighbouring farmers. The program aimed to identify and treat any animals which did not initially receive treatment for some reason during the following monthly visits to the farms. The interval between vaccinations is not critical and intervals of 2-4 months between primary and secondary immunization provide as good, or better, response to the vaccine [23]. The strategy was efficient in targeting piglets but less reliably for older animals as they were often roaming in the fields and difficult to reach.

**Fig 2.**
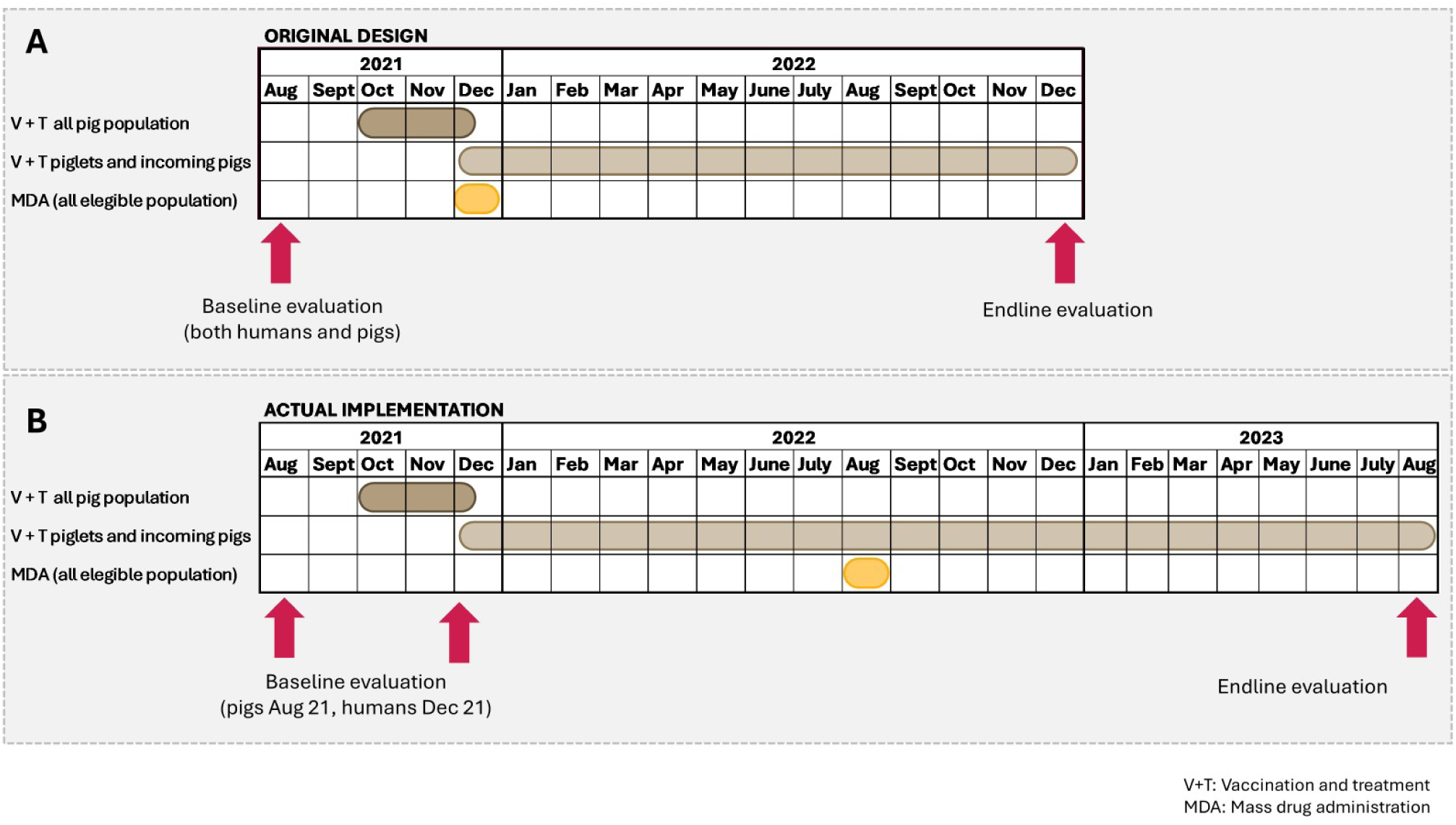
One Health model for the control of *Taenia solium* in Madagascar. Fig 2A shows the original design, and Fig 2B shows the actual implementation after the MDA had to be delayed.

Pig vaccination and treatment coverage was not measured precisely, as it would have required greater resources and logistics than were available. From August 2022, it was decided to use a proxy measure, by asking the vaccinators to record the vaccination status of the pigs they meet during their monthly vaccinations. Because the vaccinators were only going to the households where they knew there were pigs to be vaccinated (as learned from previous visits, the Chief of the fokontany or other farms), they were not capturing the total population as they were not going to households where, for example, all the pigs were already vaccinated. This is an imperfect measure as pigs could have been missed, but it helped to identify areas where the coverage was ‘lower’ and where additional awareness efforts were required.

Vaccinators were required to maintain strict biosecurity measures to prevent the spread of pig diseases, in particular African and Classical swine fevers. Disinfection of boots, overalls, lassoes, and other equipment with Virokil® was mandatory before entering and after leaving a farm. Vaccine and oxfendazole were administered using single use disposable syringes. Any waste was stored in special bags and transported to the veterinarians, who in turn disposed of it in an appropriate manner.

### Human intervention

A single MDA was undertaken in the human population once the intervention in the pig population was fully established (August 2022, Fig 2). The MDA was initially planned for December 2021, but due to adverse events with a child receiving praziquantel during a MDA program for schistosomiasis in a nearby district, the Ministry of Public Health delayed all the MDA interventions until further training and preventive measures were put in place. For that reason, the MDA was not undertaken until August 2022. The Ministry of Public Health through its communicable disease control department and the local medical staff (regional and district responsible for neglected tropical diseases, chiefs of health centers and community health agents), undertook the MDA campaign in all the eligible population over 5 years of age. The main drug used was praziquantel (Cesol™ from Merck or Biltricide® from Bayer). As MDA with praziquantel was already part of the Ministry of Public Health’s annual program for treatment of schistosomiasis (co-endemic with *Taenia solium* in this region), the MDA was conducted by the Ministry of Public Health in synergy with their MDA campaign against schistosomiasis. Children between 5 and 15 years of age targeted by the schistosomiasis program, received praziquantel at 40mg/kg. Additionally, the children in Betafo district, also received mebendazole (500mg, Vermox® from Johnson & Johnson) as part of the Ministry of Public Health program to control soil-transmitted helminths. People 15 years old and over, were treated with praziquantel at 10mg/kg as a taeniacide. Prior to the MDA, a series of training and activities were undertaken to prevent and minimise neurological adverse events (Nely et al. submitted). Eligibility criteria for praziquantel treatment in people over 5 years of age were: non pregnant women, women who had been breastfeeding for more than 3 months, and people without symptoms or signs compatible with NCC such as seizures and epilepsy or chronic headache. Those exhibiting symptoms or signs compatible with NCC were treated with niclosamide (Yomesan®, Bayer) at a dose of 1g for children from 5 to 6 years of age, and 2g for children over 6 years and adults.

Active and passive surveillance were conducted to monitor and quickly identify and manage any possible adverse events in people who had participated in the MDA (Nely et al. submitted). Local staff were trained and appropriate medicines were made available at health centers to deal with side effects, such as neurological events, drug allergies, etc.

### Training, monitoring and auditing

Prior to the interventions, training was undertaken for those involved in activities related to animals or humans.

Veterinarians and vaccinators were trained in vaccination procedures, maintenance of cold chain, identification of vaccinated animals, oral drug treatment, coverage data collection, and biosecurity measures. Messages and attitudes to transmit to farmers who may have been initially reluctant to take part of the campaign were also covered during the training. During the vaccination campaign, vaccinators were supervised and audited by local veterinarians as well as by FOFIFA staff.

Those involved in the human intervention received cascade training. Modules covered a refresher on the disease complex, standard operating procedures for the MDA (which is already part of the annual Ministry program), key messages to increase participation, coverage data collection, active surveillance of NCC cases and potential adverse reactions, and management of adverse events (Nely et al., submitted). Monitoring and supervision of activities were undertaken by the program’s senior staff pre and post-MDA campaign.

### Community sensitization

Social mobilization was a key component for this program. The local authorities were the first ones to be involved, and with their cooperation awareness campaigns were undertaken to facilitate access to beneficiaries and maximize their participation either in the animal or human interventions. Traditional singers were recruited to compose awareness-raising songs related to the fight against *T. solium* and to perform the songs in local tours in the municipalities covered by the program. In addition, radio spots were strategically broadcasted on local channels. The key messages for these two types of sensitizations were related to the parasite’s transmission, the importance of the fight against *T. solium*, encouragement of farmers to participate in the pig vaccination, encouragement of people to participate in the MDA, and the clarification of any negative rumours circulating about the interventions.

Posters were produced in local language and posted in the villages or used at community meetings. The program used the WHO/WOAH/FAO *T. solium* poster on the cycle of the parasite and the possible ways of control [24]. Other posters on the identification and management of adverse events were also prepared and used during the MDA (Nely et al, submitted).

### Data collection

Data from all interventions were recorded through paper-based forms. For pig interventions, data was recorded monthly by vaccinators and focused on the number of animals in each farm, the number of vaccinated animals according to the number of immunizations provided, and the number of non-vaccinated animals per category (< 2 months, pregnant sow, lactating sow). Local supervising veterinarians collected all data recorded and made a monthly report for the senior staff.

For the MDA, data collected related to the demographic characteristic of the person who received the drug, the presence of signs compatible with NCC, the drug administered, adverse events, the clinical signs linked to the adverse event, and the treatment received.

### Program evaluation

The program was evaluated by determining changes in the prevalence of porcine cysticercosis and the prevalence of human *T. solium* taeniasis. The objective was always to conduct the endline evaluation one year after the MDA, so because the MDA was delayed, the final evaluation was also delayed. Baseline and endline evaluations were conducted, respectively, in August 2021 and August 2023 for porcine cysticercosis. Evaluations for *T. solium* taeniasis were carried in October 2021 (repeated in December 2021) and August 2023.

Porcine cysticercosis was determined by detailed necropsy examination on 104 slaughter-age pigs selected randomly and purchased from the population. The same 26 fokontany randomly selected for sampling at baseline were also sampled at endline evaluation. The sample size for necropsy assessment (104 animals) was calculated from a population of 45,000 pigs (as per the latest official census) distributed in 20 villages, each ranging from 350 animals upwards per village, with the samples assumed to be aggregated. The expected initial necropsy prevalence was 15%. Samples size was calculated with a power of 90% and 99% confidence interval. Calculations were done in R (version 3.6.0) via Monte-Carlo simulations. Necropsies were undertaken as previously described [25]. Briefly, the animals were brought to a central veterinary facility in Antananarivo where local butchers prepared the carcasses. The muscles of the right-hand side of the carcase as well as the masseters, tongue, full diaphragm and heart were dissected from the bodies and sliced by hand at approximately 3mm intervals to reveal all cysts. Where no cysts were identified, the left-hand side of the carcase was assessed for possible infection in a similar way. Cysts were recorded as either viable or non-viable and their numbers were counted to determine the level of infection. For the endline assessments, 104 animals were randomly selected (independently of their vaccination and treatment status), and an additional 16 slaughter-age pigs were selected at random from those in the population that had not received any vaccination or oxfendazole, to have a total of 50 non vaccinated or treated pigs.

The prevalence of human *T. solium* taeniasis was determined by Kato-Katz thick smear as described in the World Health Organization (WHO) Bench aid for the diagnosis of intestinal procedures [26]. The first evaluation conducted in October 2021 only found four positive samples, which was not congruent with the high level of infection in pigs determined during the baseline necropsies. Therefore, the sampling design was adjusted, and a new purposive sampling was conducted, targeting only villages in which roaming pigs and deficient sanitation were present, but randomly selecting the people within those villages. The number of samples per village was also proportional to the number of pigs in the village, and not to the number of people, as due to the cycle of the parasite, it is less likely that there is active transmission in the larger towns in which there are fewer (or no) roaming pigs and better sanitation. In summary, for the new baseline and the final evaluation, faecal samples from 960 people were purposively sampled (according to the size of the pig population) from throughout 26 fokontany. For the baseline evaluation, one smear was prepared for most of the samples, while 2 smears were examined for 78 samples. Confirmation of the *Taenia* species present in Kato-Katz positive samples was performed by PCR and sequencing of a segment of the small subunit of mitochondrial ribosomal RNA (rrnS) gene, as described in Lightowlers et al. [27]. All endline examinations were undertaken on duplicate smears. Sample size was calculated from an estimated target population of 190,000 which would detect a reduction in prevalence from 1.8% to <0.1%, with a power of 90% and confidence level of 99%. Calculations were done in R via Monte-Carlo simulations.

### Data Analysis

All collected data were entered into Microsoft Office Excel ® spreadsheets. Statistical analyses were performed using R 4.3.0 software.

The coverage rate for pig interventions were calculated based on the number of pigs that received one dose or two doses of Cysvax and the total number of eligible pigs in the area. Pigs that were considered not to pose a risk for *T. solium* transmission were those that were less than 2 months of age, those that had received one treatment (vaccine and oxfendazole) within the previous 2 months and those that had received two treatments (vaccine and oxfendazole).

Levels of porcine cysticercosis in slaughter-age pigs in the intervention area before the intervention and in non-vaccinated or treated animals at the time of the final evaluation were compared using a negative binomial general linear model.

For human interventions, the coverage was calculated based on the number of people who received treatment during the MDA and the number of eligible targeted population. Categorical outcomes, which are the prevalences of porcine cysticercosis with viable cysts and human taeniasis before and after the interventions were compared between study groups using Chi square test /Fisher’s Exact test. A p value <0.05 was considered statistically significant.

## Results

### 3.1 Initial pig census

The number of pigs in the intervention area was assessed to be 27,071 belonging to 12,006 pig farmers, representing an estimated average of 2 pigs per farm. The majority of farmers allowed their pigs to roam freely. Some pigs went with the farmers to the field during their agricultural activities, while others roam around the villages to search for food. Most of the time, breeding pigs and their piglets, as well as pigs entering the fattening phase, are left free to roam, and some will only be confined in a pigsty at the end of the fattening phase.

### 3.2 Pig treatment and vaccination

The initial vaccination and treatment of the pig population (from October to November 2021) reached 19,135 pigs for the first immunization, representing 70.68% of the pig population determined by the census prior to the beginning of vaccinations. For the second immunizations the number of pigs vaccinated decreased to 8,120 of those previously vaccinated (p<0.001 95% CI 0.32 -0.52). This change was associated with the appearance or unsubstantiated and often outrageous rumours. Efforts were made subsequently to identify and vaccinate those pigs that missed the second scheduled immunization during the following visits, however as these animals were typically in the fields and hence unavailable when the vaccinators visited the farms.

Vaccinations that subsequently targeted piglets > 2 months and new imported pigs increased progressively in number throughout the intervention period (Fig 3). On average, 3,309 (SD ± 1,434) vaccinations per month were carried out. The number of vaccinations per month increased fourfold between January 2022 and August 2023. In total, 96,735 vaccinations were delivered during the program.

**Fig 3:**
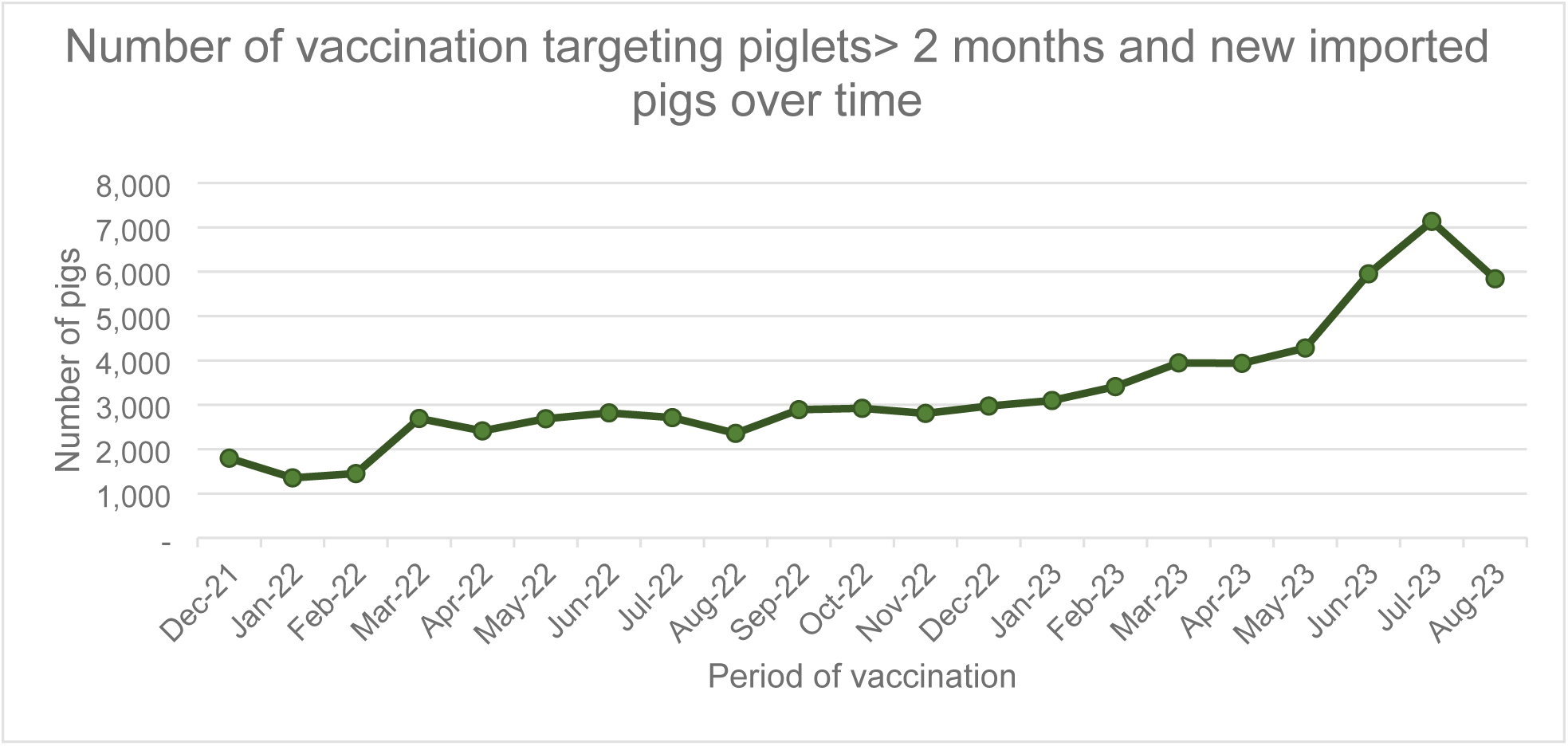
Number of pig vaccination and treatments recorded per month during the *T. solium* intervention program. Piglets>2 months and any pigs imported into the program area were targeted for vaccination and treatment, twice each, throughout the intervention period.

Data on the coverage rate of vaccination and treatments was obtained from vaccinators from August 2022 onwards and also from the random selection of pigs used for the endline evaluation necropsies.

On average, vaccinators identified approximately 7,000 pigs (eligible and ineligible, vaccinated and unvaccinated) per month. The coverage per month and the proportion of the total pig population that were considered to pose no risk for transmission of *T. solium* varied from one fokontany to another. Detailed records of vaccination coverage show a progressive increase in the percentage of the 84 fokontany achieving a coverage over 75% with >92% coverage achieved in the final months of the program (Fig 4).

**Fig 4.**
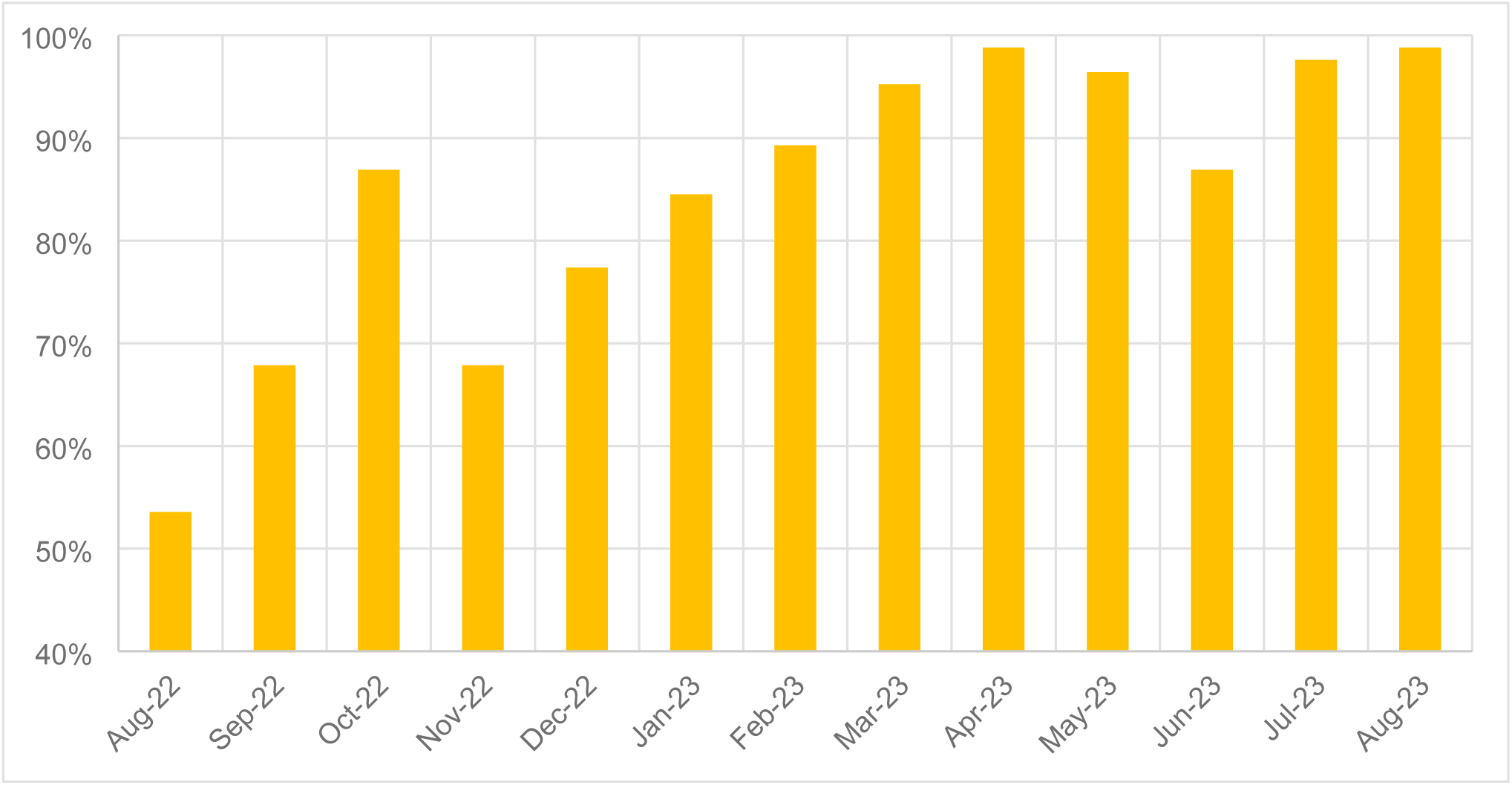
Coverage of pig vaccination and treatment over time. Proportion of fokontany that achieved a coverage ≥75% of the pig population for vaccination and treatment as recorded by the pig vaccinators.

From August 2022 when proxy detailed coverage was obtained until the end of the year (5 months), 11 fokontany out of the 84 had a coverage <50% (seven fokontany on one occasion, three fokontany on two occasions, and one fokontany on 3 occasions). In comparison, during the 8 months of 2023, only one fokontany had once a coverage below 50%.

The data for vaccination coverage from the random selection of pigs during the endline evaluation was 67.30%, which indicated that among 104 randomly selected pigs, 70 pigs were vaccinated and 34 of pigs were not vaccinated.

### 3.3 Human intervention

In total, 117,216 persons were treated during the MDA, which represented 62.52% of the eligible population (187,490 out of 231,755 total population). Significant differences in treatment coverage rates were observed between municipalities (Table 1). Fidirana municipality had the highest number of people treated, with a coverage rate approaching 90%, while Soavina represented the area with the lowest participation, with 40% of the population. There was no significant difference observed between the proportion of children under 15 treated and the proportion of people over 15 years of age that participated in the MDA (p>0.05). Among the persons treated, 615 received niclosamide as they were identified as having symptoms or signs compatible with NCC, which represent 0.52% of the treated population. Among persons treated with niclosamide, the number of people more than 15 years old was higher than the number of children less than 15 years old (p<0,0001).

**Table 1.** Population treatment data for the taeniacide MDA intervention.

| Municipalities areas | Total<br>Populati<br>on | Children<br>5 to 14<br>years old | People<br>> 15<br>years<br>old | Children 5 to 14 years old<br>treated (%) |  | People > 15 years old treated<br>(%) |  | MDA coverage<br>(%) |
| --- | --- | --- | --- | --- | --- | --- | --- | --- |
|  |  |  |  | Praziquantel | Niclosamide | Praziquantel | Niclosamide |  |
| **Ambohimasina** | 26,727 | 7,350 | 14,272 | 4,443 (40.48) | 6 (0.05) | 6,489 (59.13) | 37 (0.34) | 10,975(50.76)<sup>a</sup> |
| **Inanantonana** | 17,012 | 4,678 | 9,084 | 3,357 (33.44) | 0 | 6,681 (66.56) | 0 | 10,038 (72.94)<sup>b</sup> |
| **Ambohimanambola** | 31,037 | 8,535 | 16,574 | 5,633 (37.12) | 7 (0.05) | 9,423 (62.09) | 113 (0.74) | 15,176 (60.44)<sup>c</sup> |
| **Soavina** | 19,046 | 5,238 | 10,171 | 2,723 (43.92) | 1 (0.02) | 3,458 (55.77) | 18 (0.29) | 6,200 (40.24)<sup>d</sup> |
| **Antohobe** | 17,877 | 4,917 | 9,546 | 3,217 (39.93) | 15 (0.19) | 4,790 (59.46) | 34 (0.42) | 8,056 (55.70)<sup>e</sup> |
| **Ankazomiriotra** | 41,800 | 11,495 | 22,322 | 6,440 (36.17) | 34 (0.19) | 11,217 (63.00) | 113 (0.63) | 17,804 (52.65)<sup>f</sup> |
| **Betsohana** | 12,188 | 3,352 | 6,508 | 2,779 (32.06) | 17 (0.20) | 5,823 (67.18) | 49 (0.57) | 8,668 (87.91)<sup>g</sup> |
| **Fidirana** | 38,308 | 10,534 | 20,456 | 9,641 (34.65) | 20 (0.07) | 18,101 (65.05) | 65 (0.23) | 27,827 (89.79)<sup>h</sup> |
| **Vinany** | 27,760 | 7,634 | 14,824 | 4,205 (33.72) | 10 (0.08) | 8,181 (65.59) | 76 (0.61) | 12,472 (55.53)<sup>i</sup> |
| **TOTAL** | 231,755 | 63,733 | 123,757 | 42,438 (36.20) | 110 (0.09)<sup>j</sup> | 74,163 (63.27) | 505 (0.43)<sup>k</sup> | 117,216 (62.52) |
|  |  |  |  | 42,548 (66.76) |  | 77,668 (60.33) |  |  |

There was one case of a severe neurological adverse event. A 13-year-old child among those treated with praziquantel, presented in the evening of the first day of MDA headache, which became stronger during the night; the next day, he vomited and had seizures. This severe adverse event was promptly identified and managed as per established protocols (Nely et al submitted). A CT scan revealed three parenchymal NCC lesions. The child received appropriate medical care, there have been no recurrence of seizures and is leading a normal life.

### 3.4 Program evaluation - pigs

The randomly selected slaughter-age animals assessed at the endline necropsies comprised 70 vaccinated and treated pigs and 34 untreated pigs (Table 2). Among the vaccinated animals, no viable *T. solium* cysts were found. Five treated and vaccinated animals were found to have non-viable lesions in the muscles (4 animals with up to 10 lesions, one with 83 lesions). Among the slaughter-age pigs that had not been vaccinated and treated, 8 (23.5%) were found to be infected with viable *T. solium* cysts. Comparison of the prevalence of viable *T. solium* infection between baseline and final showed a significant difference in the prevalence of porcine cysticercosis (p<0.001). Comparison of the entire pig population between baseline and final evaluations showed a statistically significant decrease in viable infections with *T. solium* from 30.77 to 7.69%.

**Table 2.**
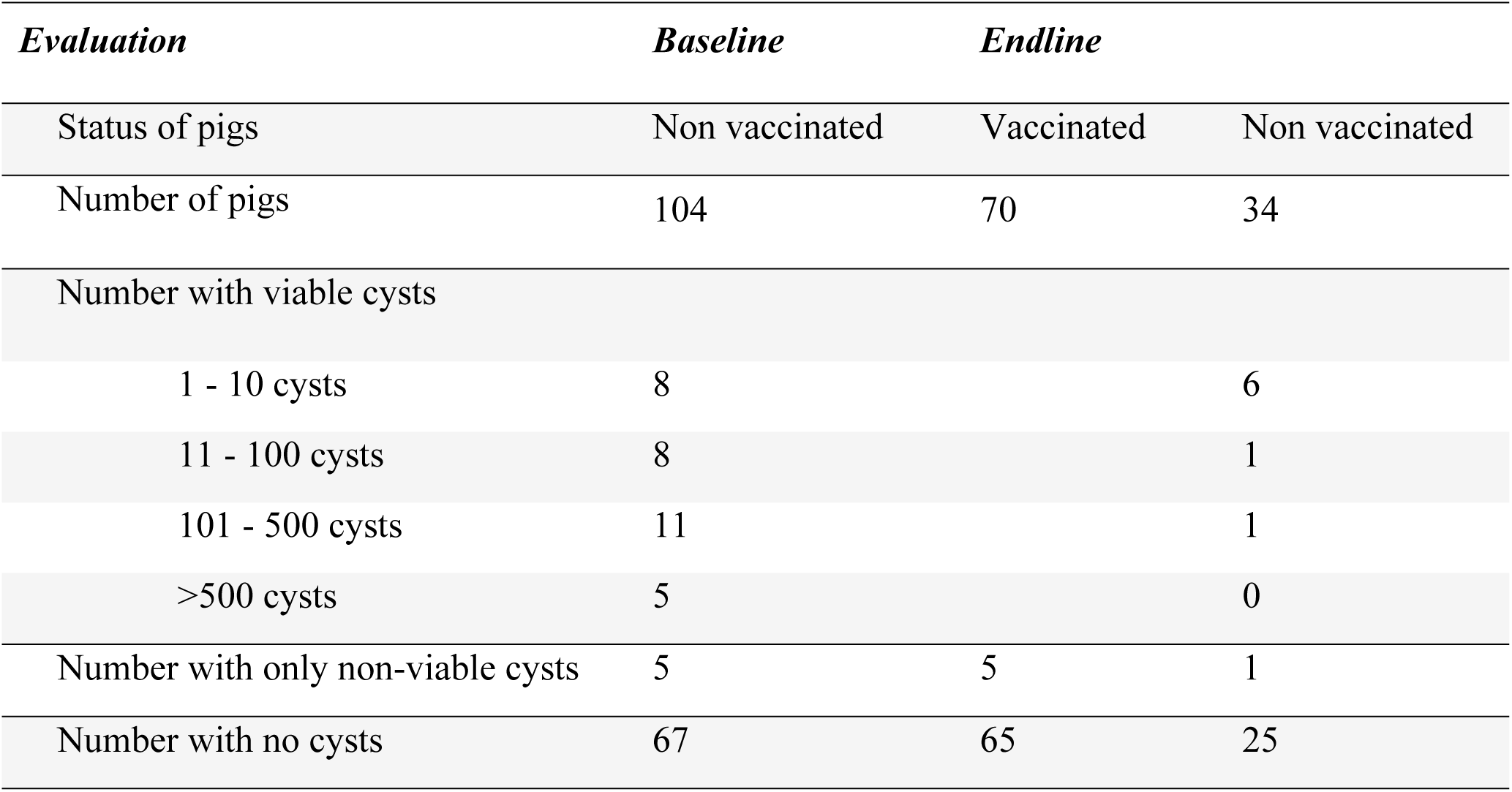
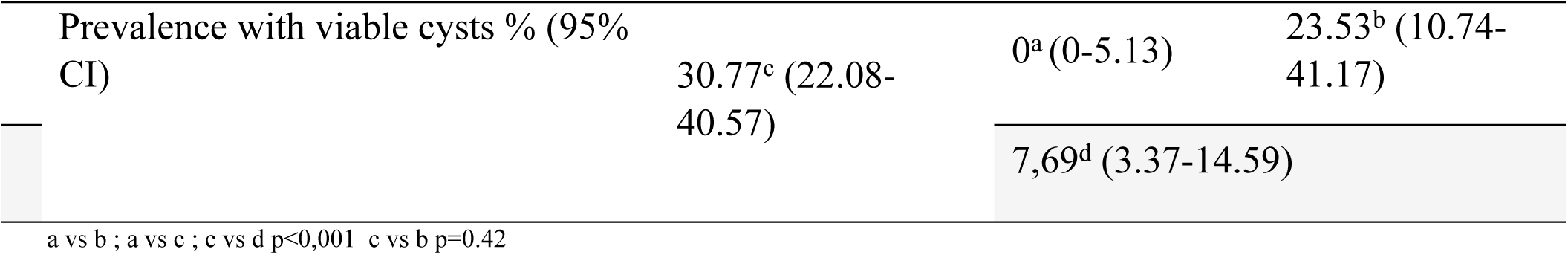
Necropsy results of the baseline and endline program evaluations.

Among the total of 50 slaughter-age pigs that were evaluated by necropsy and had not received any vaccination or oxfendazole, that is the 34 pigs randomly selected for the final evaluation plus the additional 16 randomly selected amongst the non-vaccinated or treated, 15 (30%) were found to have *T. solium* cysticerci in their muscles (either viable or non-viable).

### 3.4 Program evaluation - humans

Assessment of 960 people randomly selected from the population, Kato-Katz identified taeniasis in 12 persons (1.25%) at baseline [27] and 6 persons (0.6%) one year after the single MDA intervention. The difference was not statistically significant (p=0.13). Of the total 18 positive fecal samples, 15 (9 from baseline, and 6 from final evaluation) were able to be confirmed to species level by copro-PCR and DNA sequencing; all were confirmed as *T. solium*.

Among the samples tested at baseline, all the egg-positive samples were identified to be positive among the samples examined in a single Kato Katz slide except for one, which was identified among the 78 baseline samples tested by 2 slides, and it was positive in both slides. The six samples identified as Kato-Katz positive during the final evaluations were all positive in both slides tested.

## Discussion

The strategy of piglet vaccination and oxfendazole treatment eliminated the potential for *T. solium* transmission by the pigs that participated in the program. Of 70 treated, slaughter-age animals that were examined by slicing the entire striated musculature of the animals, no infection with viable *T. solium* was identified in any animal. Five animals were identified with non-viable cysts only. The *T. solium* vaccine targets an antigen uniquely associated with the oncosphere life cycle stage [28] and is not believed to affect cysticerci that may have established in the tissues prior to vaccination. However, the simultaneous treatment with oxfendazole kills any such pre-existing cysts in the muscles [29, 30]. For this reason, the five animals detected with only non-viable lesions in vaccinated and treated pigs at the endline necropsies are interpreted as being animals that had been exposed to infection prior to treatment [29, 30]. The important point is that these non-viable lesions present no risk of transmission of infection. By comparison, prior to the interventions 30.8% of similarly aged animals had viable *T. solium* infection in their musculature. The finding that some of the piglets were likely to have been exposed to *T. solium* prior to being vaccinated and treated highlights the importance of delivering the oxfendazole dose in full to the piglets; failure to do so could potentially result in the presence of viable cysts in vaccinated animals (due to infections established prior to vaccination) which could appear as vaccine failures.

A highly significant reduction was observed in the number of *T. solium* cysts (viable or non-viable) in non-vaccinated or treated animals that were evaluated at the final evaluation, compared to the animals assessed at baseline (Table 3, p = 0.0005). This is interpreted as reflecting a reduction in environmental contamination with *T. solium* eggs in the intervention area.

**Table 3.**
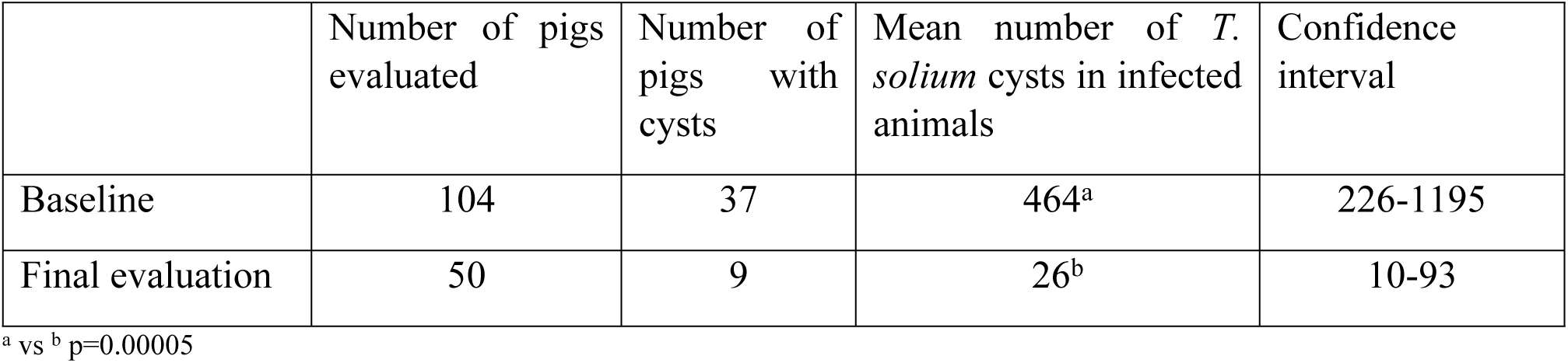
Reduction in environmental contamination with *T. solium* measured as a proxy in non-vaccinated pigs. The number of *T. solium* cysticerci (viable and non-viable) was evaluated by necropsy in non-vaccinated or treated pigs before and after the intervention.

|  | Number of pigs evaluated | Number of pigs with cysts | Mean number of *T. solium* cysts in infected animals | Confidence interval |
| --- | --- | --- | --- | --- |
| Baseline | 104 | 37 | 464<sup>a</sup> | 226-1195 |
| Final evaluation | 50 | 9 | 26<sup>b</sup> | 10-93 |

<sup>a</sup> vs <sup>b</sup> p=0.00005

There was a seasonal variation in the coverage of pig treatments, influenced by other farming activities which resulted in the farmers leaving the house very early and not being available to facilitate the vaccination, or by the farmers taking the pigs with them to the fields, such as during the months of December and January which is the rice transplanting season, or harvesting time in June. Throughout the program the coverage rate of the pig population for treatments steadily increased (Fig 4). Soon after the program began, rumours began to circulate that caused many farmers to decline to have their animals vaccinated. For example, it was believed by some that the program was a conspiracy by the government to stop the farmers from breeding local breeds of pigs. It also coincided with a high anti-vaccine general attitude in the population due to the ongoing issues concerning human vaccination for COVID. The farmers rarely vaccinated their animals for any purpose even though epidemics of Classical Swine Fever cause mass pig mortality, and hence they have a rudimentary understanding of vaccines, if any. Our use of ear tags to indicated vaccinated animals was initially also a source of concern for the farmers as it was not usual. Continuing efforts throughout the program to educate the farmers and dispel the rumours saw the dissipation of negative rumours and a steadily increasing rate of participation. The fact that treated animals were often judged by farmers to be growing faster helped improve the participation rate. This could be due to the effect of the oxfendazole treatment on nematode and other parasitic infections [31, 32].

The endline assessment in pigs was undertaken in slaughter age animals, more than 8 months of age and generally around 12 months of age (born between September 2022 and January 2023). These animals were raised when coverage of the pig population was 67% (as judged by the randomly selected slaughter-age animals taken for endline necropsy). Hence the 75% decrease in the prevalence of viable *T. solium* infection in the pig population (30.8% to 7.7%) was achieved in animals born between September 2022 and January 2023 when coverage was around 67%. Given that the vaccination and treatment of piglets eliminated *T. solium* transmission at the time of slaughter-age in all 70 animals that were assessed and had participated in the program, it could be expected that the program would have achieved a 90% reduction in transmission through pigs had it been possible to undertake necropsies at slaughter-age for animals born later in the program when the piglet coverage rate had increased to 90%. A program of continued piglet vaccination and treatment and similar intervention of animals imported into the program area, would maintain control of *T. solium* transmission indefinitely. In Madagascar it may be possible to eliminate transmission by extending the program to the entire country.

The intervention program met a major challenge with implementing the planned MDA in humans. The plan was to undertake the MDA a short time after the entire pig population had been vaccinated and treated, and piglet vaccinations were well established and continuing. However, a serious adverse event in 2021 during the routine Malagasy schistosomiasis MDA program with praziquantel resulted in a child’s death in a district nearby the program area, delaying the MDA of this program until further measures were put in place. This situation also had an impact on the population’s participation in the program’s MDA. The population coverage for the MDA was 62.5% while the objective was to reach >75% of the population. This being the case, it was not surprising to find that *T. solium* taeniasis remained present in the population at the endline assessments and transmission continued to non-treated pigs (as evidenced by the rate of infection in the 50 non vaccinated pigs). An earlier taeniasis MDA program in Madagascar, in a nearby area undertaken prior to the occurrence of the aforementioned adverse event achieved a 95% coverage [9], suggesting that the population is not averse to participating in MDA programs. Had there been more time between the adverse event that occurred during the schistosomiasis program and our MDA, a higher coverage of the population is likely to have been achieved.

An unanticipated but valuable outcome of our program has been the development of protocols, resources and a strategy to minimize the potential for adverse events occurring due to MDA being undertaken with praziquantel in an area that is, or may be, endemic for *T. solium*. These resources are available through the WHO [33, 34] and the measures are discussed in full by Nely et al. (submitted).

A feature of this *T. solium* control program was that it was implemented not as a research project, but as a public health program, carried out by the two responsible ministries in Madagascar (Ministry of Public Health and Ministry of Livestock). In this way, it was implemented by existing and already responsible structures. It was provided to all the people in the control region, without cost. The involvement of the Ministries was planned with a view to appropriation and sustainability, as well as extension once the program was completed and demonstrated to be successful. Although the program was implemented as a public health program, it was evaluated as if it had been a research project so as to obtain an accurate evaluation of its impacts.

An additional outcome of the program, was an improved relationship between the Ministries and other relevant stakeholders, reflecting the benefits of strengthening the One Health systems for future collaborations and preparedness for future challenges.

The program was designed to require minimum inputs and to have a rapid impact, so as to maximize the feasibility of the program being implemented in other *T. solium* endemic regions of Madagascar, and elsewhere. With minimum inputs we refer to only one MDA and one intervention in the whole pig population, followed only by intervention in piglets and incoming new pigs. The fact that the whole pig population is only dealt with at the start at the program, and from then onwards only piglets and new incoming pigs were treated, facilitated the field work, as having to catch large roaming pigs is not an easy task. The program provided MDA treatment drugs through a donation via the WHO and the program funding supported the cost of the vaccine, oxfendazole and vaccinators who delivered to program to the animals. Without these resources from external donors it is unlikely that control of this neglected zoonosis could be undertaken in Madagascar, or elsewhere, where full transmission of the parasite occurs.

The public health program implemented here achieved the complete cessation in *T. solium* transmission by the animals involved in the program and potentially a 90% reduction in transmission within 2 years which could be accelerated by achieving higher coverage in the MDA. This reduction would be sustainable through a continuation of the treatments in piglets, which could be reduced or terminated after *T. solium* taeniasis levels no longer provided a source of new infections. A cost-benefit analysis of this program is currently underway to provide the further evidence needed for validation. In addition, in the case of Madagascar, a map of high-risk areas for *T. solium* is being prepared so as to provide decision-makers tools for any further implementation of this public good control program.

In conclusion, this minimum input One Health *T. solium* control program significantly reduced parasite transmission. A higher MDA coverage (which is known to be feasible in the program area) and 90% piglet coverage would likely deliver a near cessation in transmission, which could be sustained by continued piglet treatments. The intervention was delivered as a public health program and therefore reflected the real-life challenges when implementing these types of programs in the field, providing a possible model for future *T. solium* control programs.

## Data Availability

All relevant data is provided as part of the submitted article

## Acknowledgement

We are very grateful to the local authorities at all levels who were involved in the implementation and the sensitization of communities program for their invaluable input. The cooperation of all inhabitants and pigs farmers in the area who participated in the interventions both in animal and human side is very much appreciated. And the program couldn’t have been implemented without the hard work and support from all the health agents, community health agents, and medical staff supporting the MDA, as well as all the pig vaccinators, and veterinarians involved in the pig activities. The success of the program is a credit to them. Support is acknowledge from the Ministry of Health, the Ministry of Agriculture and Livestock and the Ministry of Scientific Research and Higher Education.

## Notes

### Competing Interest Statement

The authors have declared no competing interest.

### Funding Statement

Yes

